# JAK-STAT pathway inhibition modulates centrally sensitised default mode network hubs in rheumatoid arthritis pain

**DOI:** 10.64898/2026.07.12.26356929

**Authors:** Kristian Stefanov, Joel T. Parkinson, Flavia Sunzini, Salim Al-Wasity, Chelsea M. Kaplan, Andrew Schrepf, Eric Ichesco, David A. Porter, Graeme A. Keith, Andrew McGucken, James Brock, Norah Aldehmi, Luz Paramo-Fiscal, Aysin Tulunay-Virlan, Maxine Arnott, Tyrone Lau, Carl Goodyear, Gregor Thut, Nicholas Shenker, Iain B. McInnes, Daniel J. Clauw, Jonathan Cavangh, Neil Basu

## Abstract

Nociplastic pain represents a major burden across immune-mediated inflammatory diseases (IMIDs). It is hypothesised, but not yet demonstrated, that peripheral inflammation promotes nociplastic pain by bottom-up sensitisation of the central nervous system (CNS). In rheumatoid arthritis (RA), a prototypic IMID, we used ultra–high field (7T) brain resting-state functional MRI to evaluate whether peripherally targeted anti-inflammatory therapies alter a biomarker of bottom-up CNS sensitisation: inferior parietal lobule (IPL)–insula connectivity. In discovery and replication cohorts, JAK-STAT pathway inhibitors significantly shifted IPL–insula connectivity toward a normalised pattern. This effect was not seen with placebo or anti-TNF therapy. Moreover, after performing an agnostic whole-brain multivariate analysis of functional connectivity change related to JAK-STAT inhibition, the posterior cingulate cortex (PCC) was identified; like the IPL, a major hub of the default mode network (DMN). We then probed the DMN with transcranial magnetic stimulation in an independent RA cohort. Active, but not sham, stimulation altered DMN connectivity and reduced peripheral blood monocyte pSTAT3 function, a surrogate of immune inactivation, suggesting a bi-directional brain–immune circuit. Together, these findings provide the first human experimental evidence that peripheral JAK-STAT pathways contribute to nociplastic pain in IMIDs.

**One Sentence Summary:** Blocking peripheral JAK-STAT signalling reduces both objective and subjective nociplastic pain in rheumatoid arthritis, revealing a localised inflammatory driver behind a traditionally non-inflammatory pain state.

## INTRODUCTION

Central sensitisation comprises the maladaptive activity of nociceptive pathways in the brain and spinal cord, which primarily underpin nociplastic pain. This mechanistically distinct pain subtype defines multiple primary chronic pain conditions, including fibromyalgia (FM) - the prototypical nociplastic pain disorder. With an estimated global prevalence of 2-3%, FM is the third most frequent musculoskeletal condition(*1*). As a primary cause of loss of life quality(*2*), it represents a commanding unmet public health need by disabling individuals(*3*), but also by driving a considerable economic burden to society(*1, 4*).

There are currently no recommended nociplastic pain targeted therapies, and thus, a greater understanding of pathogenesis is essential. A role for systemic inflammation as a determinant of central sensitisation is suggested by the clinical observation that nociplastic pain co-exists disproportionately among people with immune-mediated inflammatory diseases (IMIDs). For example, rheumatoid arthritis (RA), a prototypical IMID, engenders 10 times more likelihood to fulfil FM classification criteria compared to the general population(*5*).

We and others previously hypothesised that, in the context of systemic inflammatory disease, a ‘peripheral’ inflamed tissue (that resides outside the anatomical nervous system) drives ‘bottom-up’ inflow to sensitise the central nervous system (CNS). This manifests phenotypically as FM in a significant proportion of IMID patients. Characterisation of this endotype would pave the way for entirely novel approaches to the management of pain in those with IMIDs. Specifically, strategies that directly treat the CNS combined with those that indirectly attenuate ‘bottom-up’ driven pain pathways would be beneficial for a substantial subset of patients. Critically, this was the original meaning of the term central sensitisation, wherein ongoing peripheral nociceptive input led to the CNS sensitisation in animal models(*6*).

Employing functional MRI (fMRI) of the brain, we previously identified abnormal connectivity between the default mode network (DMN) and the insula (a pronociceptive brain region) in patients with RA and coexisting nociplastic pain: a consistent biomarker of central sensitisation(*7*). We further showed that aberrant connectivity between the inferior parietal lobule (IPL), a DMN component and a sensory region linked to the neural response to systemic inflammation in RA(*8*), and the insula was associated with systemic inflammation, but only in RA patients manifesting features of nociplastic pain(*9*). Thus, IPL-insula functional connectivity separated those RA patients apparently vulnerable to ‘bottom-up’ central sensitisation.

These observational data were cross-sectional and indirect; intervention-based experimental evidence to support this ‘bottom-up’ concept has been lacking. Moreover, pain is a ‘problem of neural networks’, rather than single regions, and the wider neurobiological map of this novel explanation of central sensitisation has yet to be defined. The necessary characterisation is not limited to the brain since the peripheral-central interplay underlying this concept could constitute a homeostatic circuit, as demonstrated in the established neuroinflammatory reflex, where the CNS has a role in both monitoring and regulating systemic inflammation(*10*).

We hypothesise that peripherally directed anti-inflammatory therapies will attenuate a putative bottom-up pathway and thereby modify the subjective clinical assessment of central sensitisation and alter functional connectivity between IPL and insula. Standard RA care is based on treatments that effectively attenuate the nociceptive input of peripherally inflamed tissues, which herewith we have leveraged to test our hypothesis. These include JAK-STAT targeted therapies, which have demonstrated a notable analgesic effect in randomised controlled trials(*11*). By employing high-resolution 7T fMRI for the first time in RA, we gained greater statistical power than available using lower strength magnets to allow us to investigate the pivotal brain hubs, which subserve this construct. Thereafter, we applied repetitive transcranial magnetic stimulation (rTMS), a non-invasive neural probe(*12*), to modulate our identified brain regions of interest and interrogate a putative bi-directional relationship with systemic inflammation. This combination of phenotypic description to inform ‘region-targeted’ active perturbation is a unique approach to our knowledge. On this basis, we report, for the first time, human clinical experimental data that inform the characterisation of a novel neuro-immune circuit which subserves nociplastic pain in RA.

## RESULTS

Twenty-one RA patients (16 female; mean age, 54.1 years) with moderately to severely active disease comprised the discovery cohort (JAKiDiscovery). They were invited to attend an MRI brain scan just prior to initiation of filgotinib (JAK-1 inhibitor, 200mg daily) and at 12-week follow-up, in addition to an optional 4-week MRI brain scan. We also sought to replicate our findings in a second cohort (JAKiReplication) of thirteen moderately to severely active disease RA patients (9 female, mean age 58.0) who initiated baricitinib (JAK-1/2 inhibitor, 4mg daily). See Fig. S1 for further details.

### JAKi normalised negative IPL-insula connectivity across two RA cohorts

Our principal hypothesis was that peripherally directed anti-inflammatory therapies would alter a bottom-up component of central sensitisation in RA, as measured by functional connectivity between the left IPL and insula.

Evaluation of the left IPL–left posterior insula connectivity between baseline and 12 weeks revealed a significant change in the JAKiDiscovery (*t* (15) = -2.56, p FDR = 0.043, Cohen’s d = 0.64) cohort and replicated in the JAKiReplication (*t* (8) = -2.33, p = 0.048, Cohen’s d = 0.78) cohort. Notably, connectivity shifted toward zero (i.e. became less negative) **(Fig. 1**), indicating normalisation of this connection and reduced bottom-up drive on central pain circuits. Furthermore, this normalisation appeared as early as four weeks in the JAKiDiscovery subgroup (n = 7), who completed an optional 4-week scan (Error! Reference source not found.). This effect was not observed after anti-TNF (n=21, mean age 59.3) or placebo (n=9, mean age 51.1) in an independent, secondarily analysed, RA cohort of moderate/severe active disease, in which an identical 7T MRI protocol was acquired (Error! Reference source not found., Error! Reference source not found.).

**Fig. 1.**
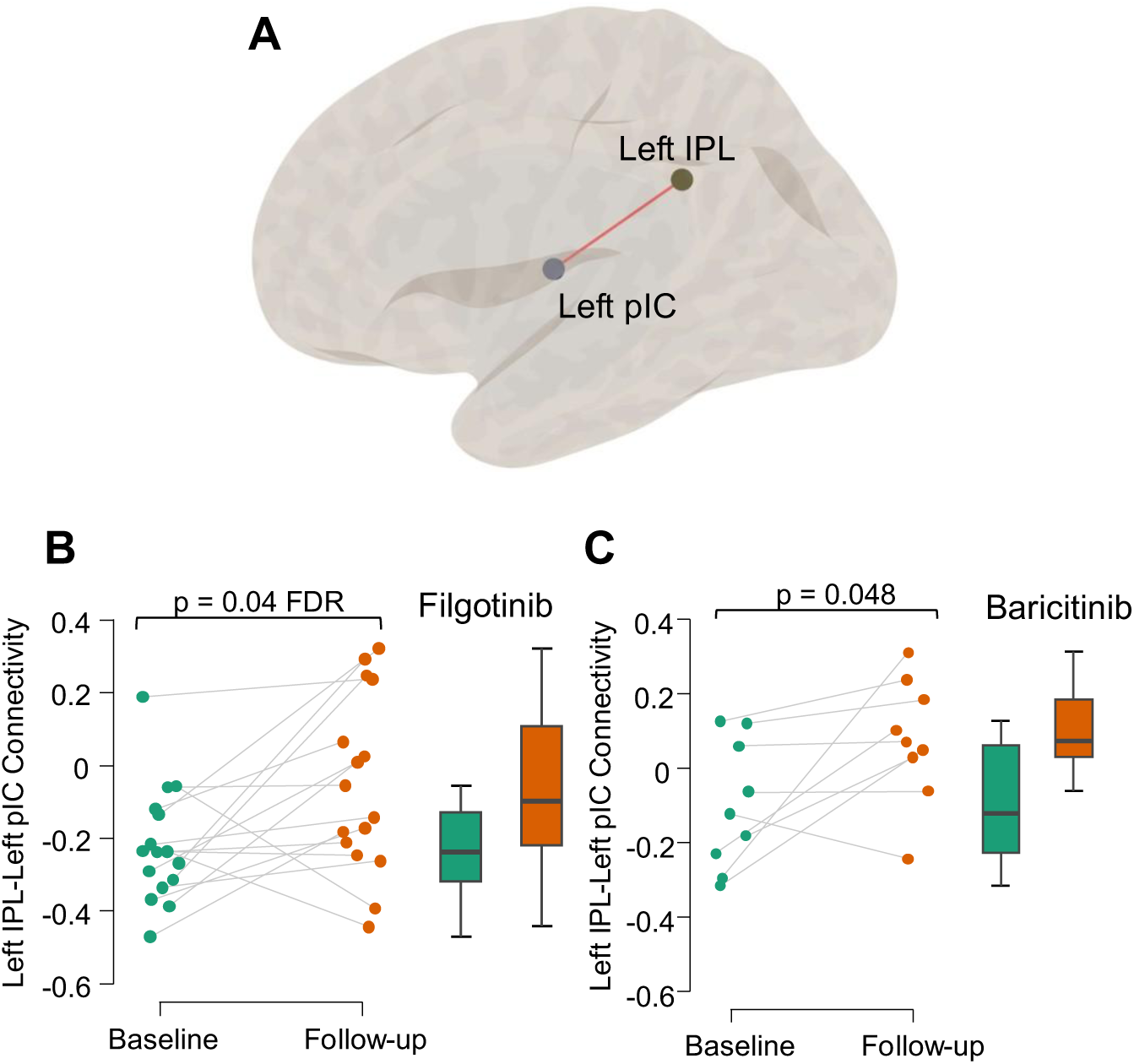
JAKi effect on left IPL with left posterior insula functional connectivity. (**A**) Lateral view of the two seeds of the left inferior parietal lobule (IPL) and the left posterior insular cortex (pIC) used in the analysis. The boxplots on the lower panel depict the significant change (paired t-tests between baseline and 12-week follow-up) in functional connectivity between the two seeds after filgotinib in the JAKiDiscovery (**B**) and baricitinib in the JAKiReplication (**C**).

### Agnostic analysis of JAKi change identifies the main hub of the DMN

To extend beyond candidate based, IPL–insula tests, we applied a whole-brain multivariate analysis (fc-MVPA) to identify additional brain hubs whose connectivity may support a neuro-immune circuit. We combined the two JAKi datasets to permit fc-MVPA, whose sensitivity can approximately double with increasing participant numbers(*13*). JAKi significantly changed the connectivity of a posterior cingulate cortex (PCC) cluster with the rest of the brain (**Fig. 2**, peak coordinates = -8, -34, 36, cluster size = 132 voxels, *F* (5,115) = 4.9, p FDR < 0.001). This cluster lay predominantly within the DMN, with 85% of its voxels overlapping Yeo’s DMN atlas(*14*). This implicates a canonical DMN hub as a candidate integrator linking immune-related changes with large-scale network reorganisation.

**Fig. 2.**
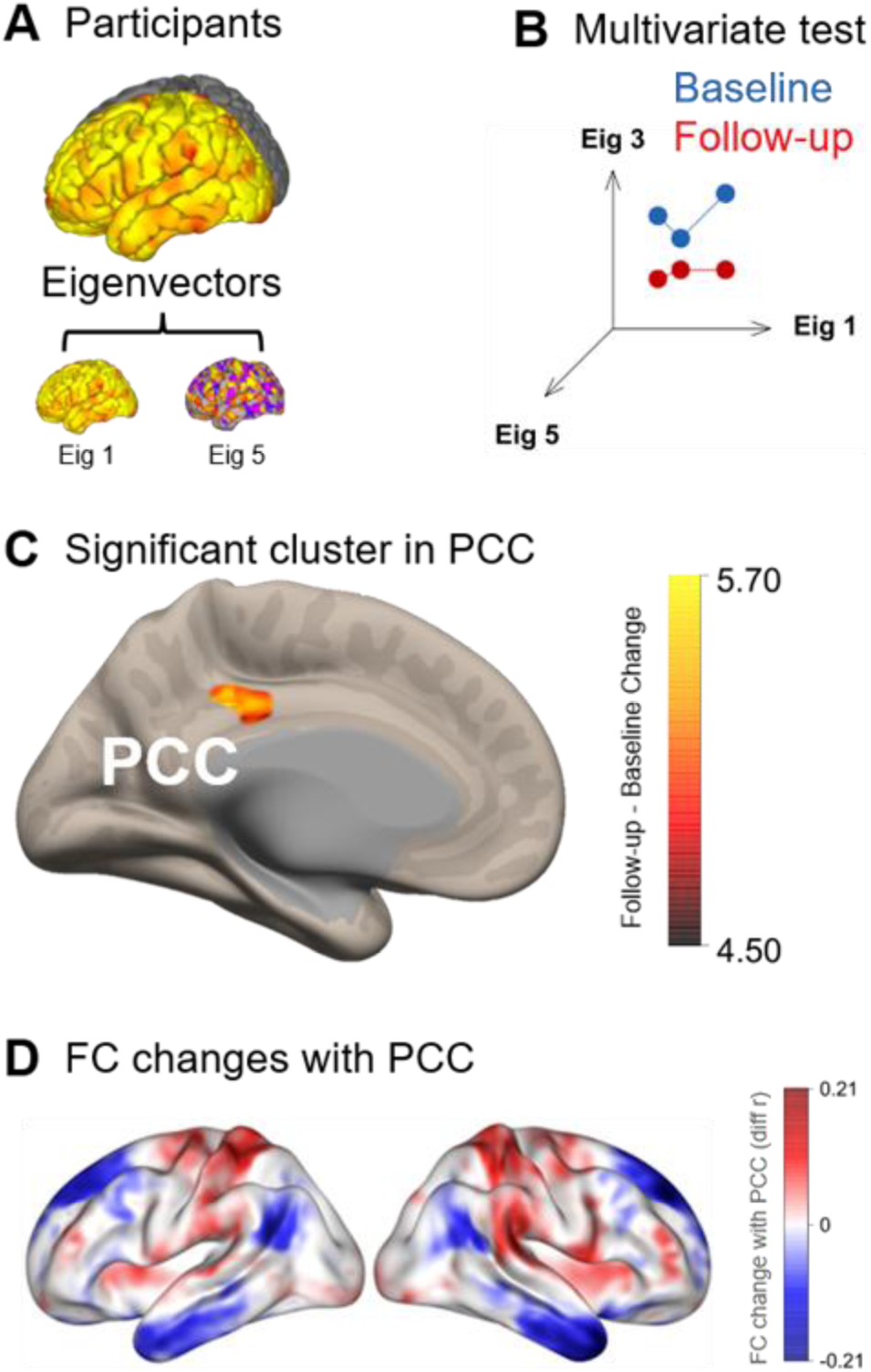
fc-MVPA in JAKi identified the PCC as significantly changed. **(A)** Five eigenvectors were extracted from baseline and follow-up functional connectivity data from participants. **(B)** A multivariate test contrasted follow-up against baseline eigenvector connectivity. **(C)** The test identified that the pattern of connectivity of a cluster in the posterior cingulate cortex (PCC) was significantly changed. **(D)** Connectivity changes between the PCC and the rest of the brain are illustrated on the left and right hemispheres (blue = reduced connectivity, red = increased connectivity).

### JAKi normalised DMN-insula connectivity

Given the apparent prominence of the IPL and PCC, we expanded our analysis to examine broader DMN connectivity with the insula (**Fig. 3A**). We compared within-participant baseline and 12-week resting-state connectivity between the DMN and bilateral insula in the combined JAKi dataset using paired t-tests (Error! Reference source not found.). We defined the DMN using Yeo’s network atlas(*14*) and extracted it in both JAKi cohorts. DMN–right anterior insula connectivity changed after treatment (*t* (23) = -2.4, p = 0.025, Cohen’s d = 0.49, **Fig. 3B**). Mean connectivity shifted from negative at baseline (−0.398, SD = 0.195) toward zero at follow-up (-0.28, SD = 0.26), mirroring the normalisation seen in left IPL–left posterior insula connectivity.

**Fig. 3.**
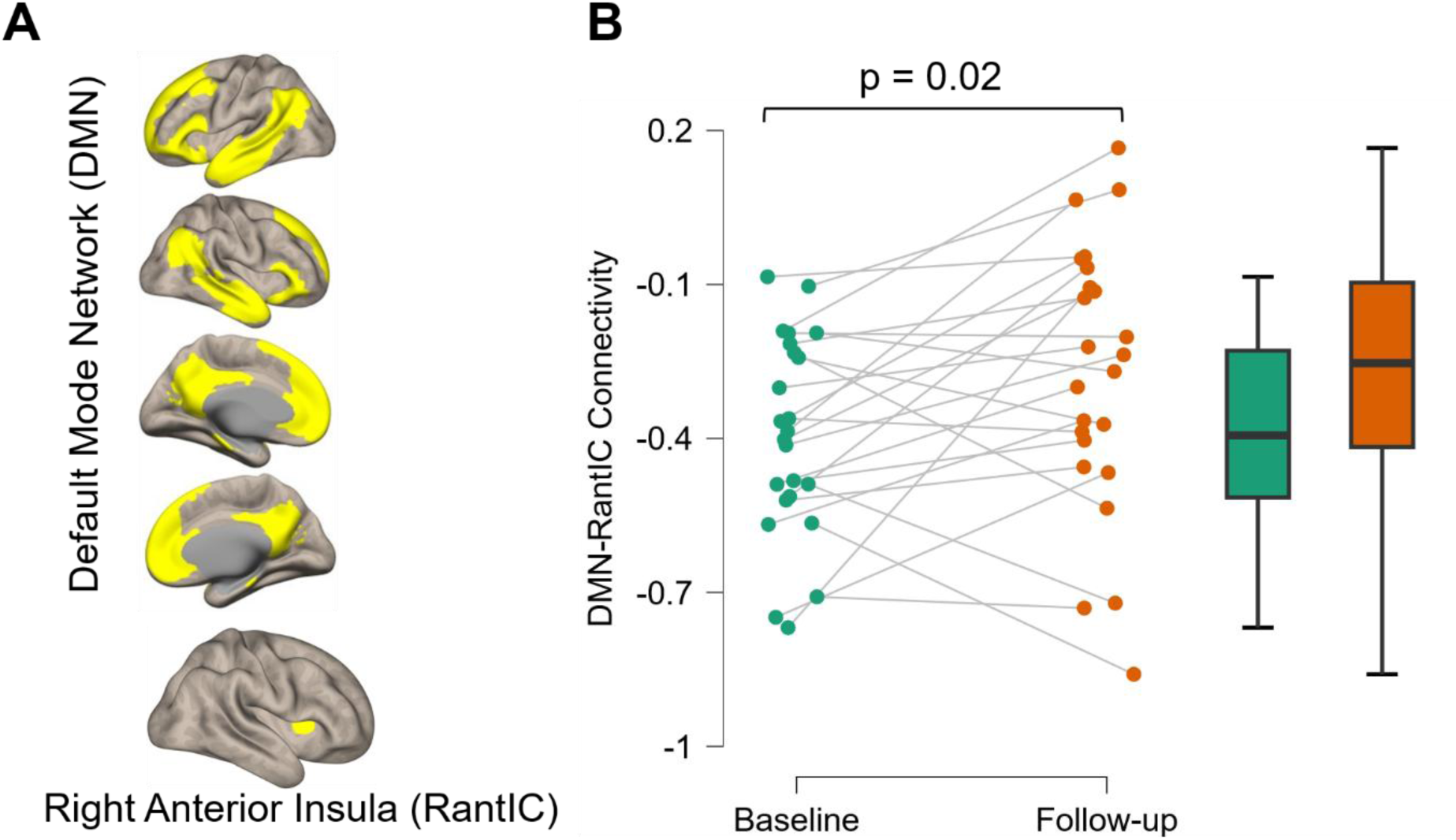
JAK inhibition normalised DMN-insula connectivity. (**A**) Visualises the right anterior insular cortex (RantIC) seed and the default mode network (DMN) based on the Yeo network atlas. (**B**) Boxplot displays the significant change (paired t-test) in DMN-RantIC connectivity in the combined cohort of patients who received either baricitinib or filgotinib.

### Clinical effects of JAKi

The subjective measure of nociplastic pain aligned with the imaging findings. In the combined JAKi cohort, a statistically significant improvement in self-reported nociplastic pain (ACR FM score; -5.48 (*t* (24) = -4.48, p = 0.001, Cohen’s d = 0.9) was reported. Further support for a putative CNS effect came from a significant reduction in depression scores (*t* (24) = -2.38, p = 0.03, Cohen’s d = 0.48). Consistent with clinical trials (*15*), JAKi improved overall disease activity. At 12 weeks, mean disease activity (DAS28) improved significantly (mean difference = -1.39, *t* (24) = -4.74, p=0.001, Cohen’s d=0.95) (**Table 1**). Curiously, however, there was no statistically significant improvement in CRP. Although underpowered, these findings suggest that the clinical effects of JAK inhibition may extend beyond suppression of classical peripheral inflammation markers alone.

**Table 1.** Clinical profile of rheumatoid arthritis patients in the combined JAKi cohort overall and by sex before and after treatment, and the statistical significance of those changes. Values depict mean (± SD) while overall statistic shows t value of paired-samples t-tests between baseline and follow-up (12 weeks) of total cohort and their respective p-values; ^ CRP had a non-normal distribution, so the table shows its median and IQR and the result of a Wilcoxon signed-rank test instead of a Student t-test with a reported z-statistic.

| Variable | Timepoint | Total cohort<br>(n=25) | Female<br>(n=17) | Male<br>(n=8) | Overall<br>statistic | p |
| --- | --- | --- | --- | --- | --- | --- |
| Age |  | 56 (10) | 56 (9) | 56 (11) |  |  |
| Disease<br>Duration |  | 13 (10) | 14 (11) | 10 (8) |  |  |
| DAS28 | Baseline | 4.62 (1.41) | 4.29 (1.27) | 5.34 (1.5) | -4.74 | <.001 |
|  | Follow-up | 3.23 (1.06) | 3.18 (1.06) | 3.35 (1.11) |  |  |
| SJC | Baseline | 7.8 (6.24) | 6.82 (6.05) | 9.88 (6.51) | -1.91 | .068 |
|  | Follow-up | 4.64 (5.89) | 3.53 (4.37) | 7 (8.12) |  |  |
| TJC | Baseline | 10.6 (8.68) | 8.94 (8.3) | 14.13 (8.95) | -4.20 | <.001 |
|  | Follow-up | 4.92 (6.81) | 4.59 (5.44) | 5.63 (9.5) |  |  |
| CRP<sup>^</sup> | Baseline | 3 (10) | 3 (5) | 8.5 (19.25) | -1.31 | .19 |
|  | Follow-up | 3 (6) | 2 (5) | 3.5 (4.75) |  |  |
| ACR FM score | Baseline | 14.12 (5.53) | 14.41 (4.64) | 13.5 (7.41) | -4.48 | <.001 |
|  | Follow-up | 8.64 (6.34) | 8.88 (5.29) | 8.13 (8.58) |  |  |
| Pain NRS | Baseline | 5.31 (2.26) | 5.18 (2.51) | 5.56 (1.81) | -5.37 | <.001 |
|  | Follow-up | 2.43 (2.25) | 2.41 (2.23) | 2.48 (2.46) |  |  |
| Depression<br>score | Baseline | 18.28 (7.99) | 18.88 (8.48) | 17 (7.17) | -2.38 | .03 |
|  | Follow-up | 14.48 (6.49) | 15.24 (6.54) | 12.88 (6.49) |  |  |

### Non-invasive brain stimulation modulated IPL-PCC connectivity

Having observed that peripherally directed (typically non-blood-brain-barrier penetrant) anti-inflammatory JAKi lead to changes in DMN brain connectivity, we next evaluated whether the DMN participates in a top-down peripheral immune effector circuit. This is a critical component of our bi-directional brain–articular axis proposition. Based on the above data, and as the most accessible DMN hub, we targeted the IPL, postulating that its perturbation would engage PCC connectivity and elicit a systemic immune response. In a further independent cohort of RA patients who were receiving stable non-JAKi medication and reported chronic pain, we applied repetitive transcranial magnetic stimulation (rTMS) to the left IPL. The rTMS was used within a randomised, sham-controlled crossover design, with each participant receiving active and sham stimulation on separate study days. We obtained blood samples and resting-state fMRI 1 hour before and after each session. Nine RA patients completed all pre/post resting-state fMRI sessions, with one patient excluded due to excessive motion. Active rTMS decreased left IPL–PCC connectivity (**Fig. 4A**, n = 8, Wilcoxon signed rank test, Z = 2.4, p = 0.02, rank biserial correlation = 0.94), whereas sham stimulation produced no measurable change (*t* (7) = 0.29, p = 0.78; **Fig. 4B**). DMN (PCC, IPL) connectivity with known neuro-immune effector regions was then documented. These included regions in the lower brain stem(*16*) and the hypothalamus(*17*). Notably, active rTMS changed connectivity between the PCC and the Locus coeruleus (*t* (7) = 3.13, p = 0.02) and between the left IPL and hypothalamus (*t* (7) = 2.33, p = 0.05) (Error! Reference source not found.).

**Fig. 4.**
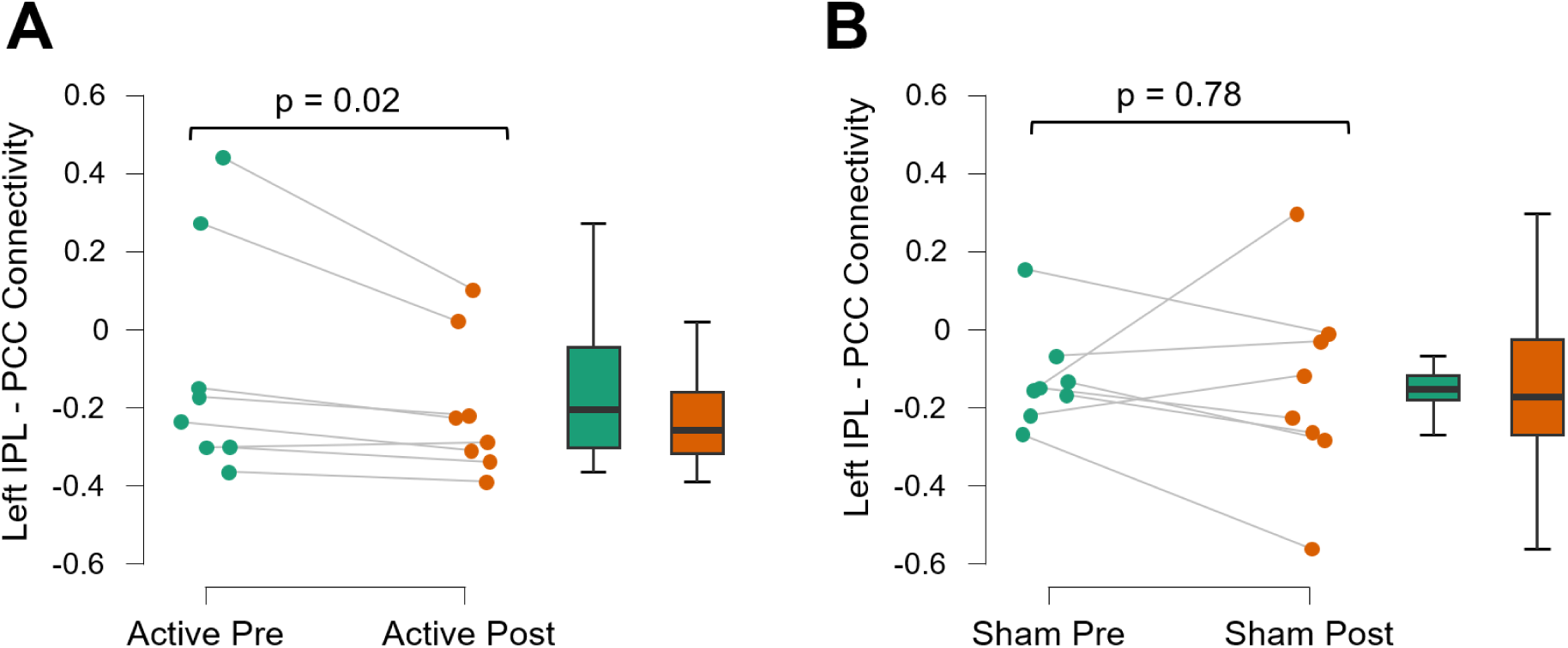
Brain modulation affects left IPL-PCC Connectivity. The boxplots depict change after active stimulation using high frequency (10Hz) repetitive transcranial magnetic stimulation (rTMS) compared to sham (control) stimulation, where a device mimicked the stimulation experience without affecting the brain. **(A)** Active rTMS significantly (Wilcoxon singed-rank test) reduced connectivity between the left inferior parietal lobule (IPL) and the posterior cingulate cortex (PCC). **(B)** This was not observed with sham stimulation (paired t-test, p = 0.78).

### Exploratory interactions between CNS modulation and peripheral blood signalling status in monocytes

We next explored the possibility that a parallel effect upon pSTAT3 levels (an established assay of JAK 1/2 inhibitor(*18*) pharmacodynamic effect) on blood monocytes might be detected. The JAK-STAT pathway comprises a putative afferent arm of our circuit, and its capacity to modify immune responses within the short time frame available (since intracellular phosphorylation levels are characterised by quicker kinetic changes compared to protein expression) enabled our experimental design. Of the eight patients included in the neuroimaging analysis, 2 were excluded from the final analysis due to poor retrieval of a monocyte population from frozen PBMCs (below 2% of live singlets PBMCs). Active rTMS of the left IPL induced a rapid decrease in pSTAT3 in monocytes (**Fig. 5**). Albeit reduction of pSTAT3 did not reach statistical significance in active rTMS (*t* (5) = 2.1, p = 0.08, Cohen’s d = 0.87), a time-by-condition interaction was present (*F* (1,20) = 5.78, p = 0.026) because sham stimulation showed no change (*t* (5) = -0.86, p = 0.43). Overall, these results suggest a brain-to-immune signalling effect (**Fig. 6**).

**Fig. 5.**
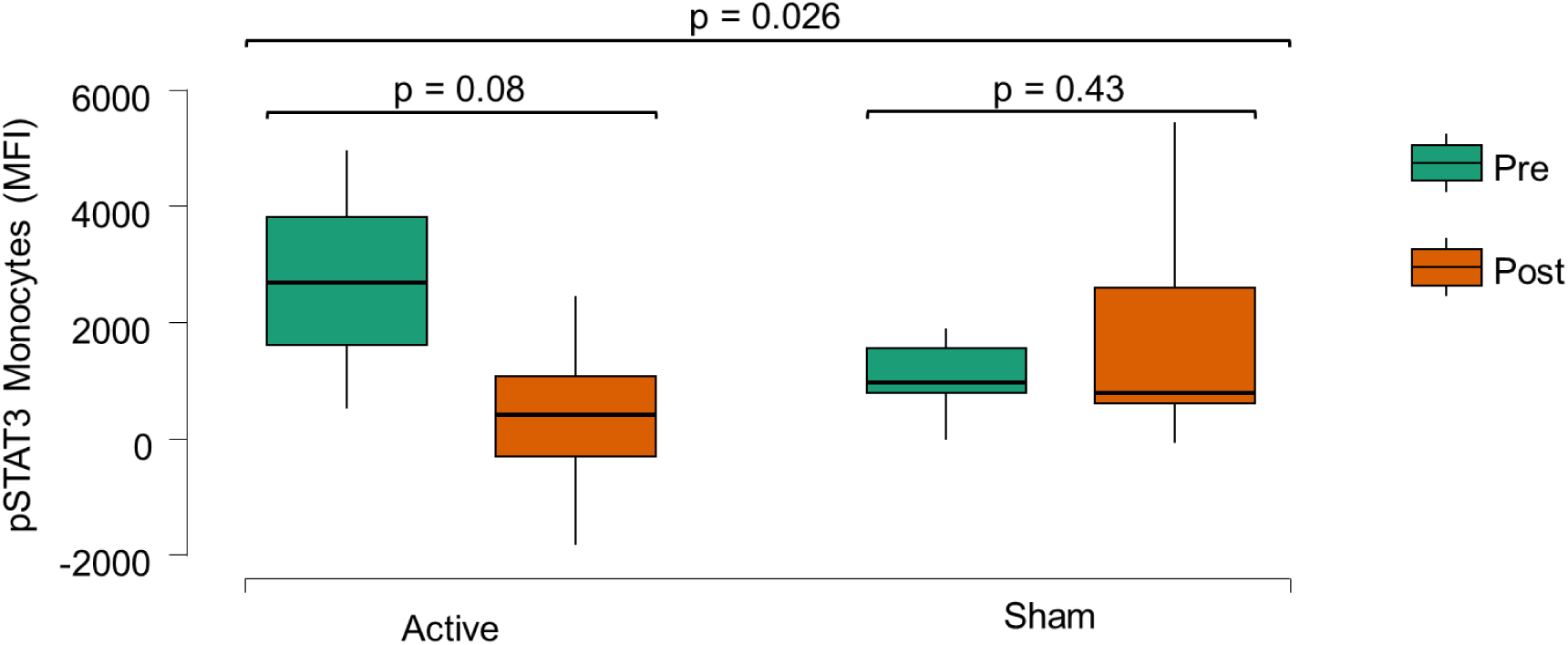
Brain modulation affects pSTAT3 in monocytes. pSTAT3 expression is reduced (trend level) in CD14+ monocytes following active rTMS modulation of the left IPL. There is a significant interaction (linear mixed model) between time (pre, post) and condition (active, sham).

**Fig. 6.**
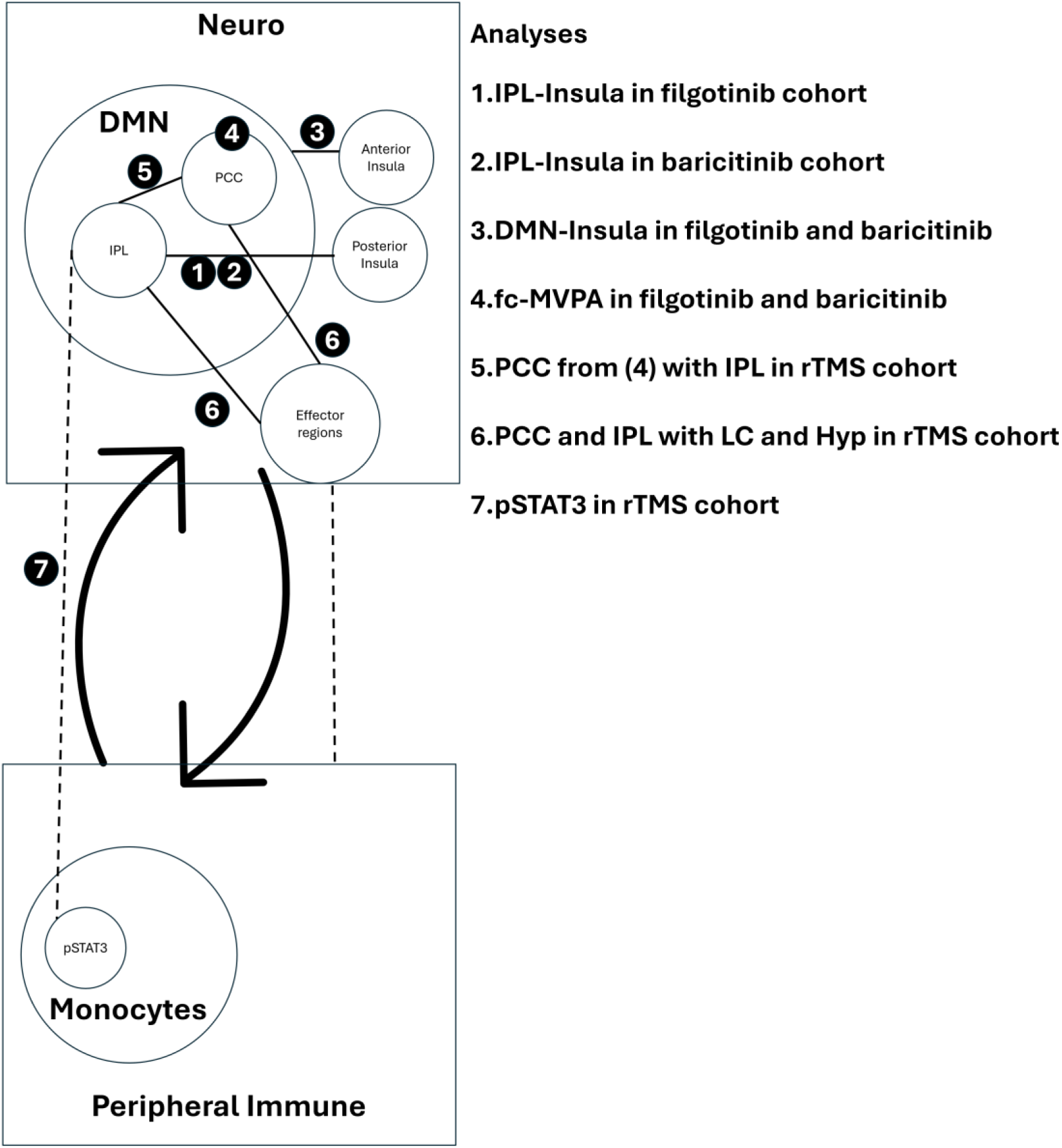
Neuro-peripheral circuit. Our observed treatment-associated shifts in IPL–insula and DMN–insula coupling and rTMS left IPL- rTMS pSTAT3 change suggest a potential bi-directional neuro-peripheral circuit underlying nociplastic pain: the IPL functions as an inflammation-responsive sensory region, the posterior insula indexes amplified signals of central sensitisation–like pain, and the PCC and IPL integrate these signals within the DMN to coordinate adaptive homeostatic responses in effector regions like Locus coeruleus (LC) and the Hypothalamus (Hyp).

## DISCUSSION

Individuals with IMIDs such as RA develop nociplastic pain at much higher rates than in the general population, and we assume that systemic inflammation may trigger and/or perpetuate the nociplastic phenotype by driving central sensitisation of the nervous system. However, this assumption has yet to be experimentally tested in humans. Here we report that people with RA not only experience significant subjective improvements in nociplastic pain following therapy with a peripherally targeted anti-inflammatory drug but also demonstrate significant objective normalisation of a neurobiological marker of ‘bottom-up’ central sensitisation: IPL to insula functional connectivity.

To the best of our knowledge, we are the first to document the responsiveness of a subjective metric of nociplastic pain to anti-inflammatory interventions in RA. These data align with a previous observational study in axial spondyloarthritis, which similarly demonstrated that such treatments predicted nociplastic pain recovery(*19*). However, subjective scales of central sensitisation have yet to be validated in inflammatory disease, and the currently available measures are unable to fully distinguish between central and peripheral origins of pain. Moreover, these measures consider central sensitisation as a single construct and fail to specifically quantify a ‘bottom-up’ subtype. Alternatively, the DMN, and specifically its connectivity with the insula (a critical hub of the salience network, SLN) as measured by fMRI, is one of the most reproducible objective markers of central sensitisation(*20*), and is distinct from brain regions which classically sense peripheral nociception, such as the somatosensory cortex. In RA, the consistently observed association between systemic inflammation and a subcomponent of this brain marker, IPL-insula, provides a proxy for ‘bottom-up’ central sensitisation, which, following our fc-MVPA analysis, may now be extended to include the PCC, another major hub of the DMN.

We observed a shift in left IPL–insula coupling from strongly negative toward zero, indicating less synchronised activity between the two regions, typical in healthy resting brain organisation. Resting-state fMRI studies in healthy subjects show that interactions between the DMN and SLN vary over time to support flexible switching between self-referential processing and attention to salient events in the environment or the body(*21*). DMN–SLN coupling is typically small and highly state-dependent(*22*), which is seen across the lifespan(*23*). In nociplastic pain, within network connectivity is decreased in the DMN but increased in the SLN, while higher pain sensitivity is linked to stronger DMN–SLN coupling between the insula and lateral parietal cortex(*24*). In our cohort, we hypothesised that JAK inhibition reduces the salience of ongoing pain, which would reduce SLN influence over the DMN, consistent with the observed change in left IPL-insula connectivity. Clinically, this rebalancing may underlie pain relief by supporting endogenous pain modulation and reducing persistent self-referential engagement with pain.

Our observations are potentially specific to JAKi since results were not replicated among anti-TNF (adalimumab) or placebo groups (Error! Reference source not found.). Clinical trial data consistently report superior analgesic effects for JAKi in comparison to anti-TNF therapies in RA(*11*). Post-hoc analyses of these data fail to fully attribute this difference to the added suppression of classical systemic inflammation(*25*). Alternative mechanisms of peripheral action have been inferred from pre-clinical experiments of pain in the collagen antibody-induced arthritis model. Greater reduction of pain behaviours and peripheral nociceptive markers (synovial sensory nerve fibre outgrowth, neuronal excitability, dorsal root ganglion glia activity) were noted among mice treated with JAKi versus anti-TNF(*26*). Adaptor protein-2 associated kinase 1 (AAK1) signalling was implicated in this study, but there is growing recognition of other putative targets, such as the stromal cell product Netrin 4, which are not considered classical inflammation mediators but appear to define pain in the RA joint(*27*). Such findings increasingly track with the common clinically observed disconnect between canonical index markers of inflammation, such as CRP, and RA pain(*28*) and are reinforced by the exploratory clinical outcomes of this study, which demonstrate significant pain reductions in the absence of significant CRP reductions. Regardless of the precise immune mediation, it is hypothesised that the common end effect of therapies, such as JAKi, is peripheral, with subsequent orchestration of central sensitisation.

An alternative, or co-existing, explanation is that JAK inhibitors influence the CNS more directly via the blood compartment. Unlike anti-TNF therapies, which are large monoclonal antibodies (>140,00 Da)(*29*), JAK inhibitors are small molecules (>500Da) that theoretically could penetrate the blood-brain barrier (BBB). Robust evidence supporting this possibility is, however, constrained to pre-clinical studies, including a series of collagen-induced arthritis experiments which demonstrated that peripheral administration of baricitinib attenuated microglial activation in the area postrema, a region of the brain with relatively weakened BBB function(*30*). Although we consistently demonstrated brain alterations subsequent to JAKi therapy, it cannot be determined whether these are direct CNS effects or indirect effects driven by peripheral signalling. We also interpret the differential effects with anti-TNF and placebo cautiously since these were secondary analyses of separately recruited cohorts. Although the eligibility criteria were similar for these parallel studies, baseline clinical measures and fMRI profiles differed (see Error! Reference source not found.), including weaker IPL–insula connectivity. This weaker baseline coupling suggests more modest “bottom-up” sensitisation and less scope for “normalisation”. While this prohibits absolute proof of JAKi specificity, it does motivate future research into the use of IPL-insula functional connectivity as a stratification marker and studies to evaluate its performance in identifying those RA patients with a co-existing FM phenotype whose pain is most likely to benefit from systemic inflammation targeting.

Although our primary objective was to determine ascending systemic immune-CNS communication, bi-directional relationships are inherent to most complex physiological phenomena. For example, in RA, the neural inflammatory reflex—comprising vagal sensors of pro-inflammatory cytokines such as TNF-α and IL-1β, brainstem integrators in the nucleus tractus solitarius, and splenic nerve effectors of myeloid function (*31*)—has been successfully targeted with vagal nerve stimulation (VNS), which is now FDA-approved for the management of RA disease activity(*32*). We therefore explored the possibility of a different neuro-immune circuit by employing TMS to modulate our putative CNS sensors/integrators of peripheral inflammation to determine any descending CNS-peripheral immune effect. We examined pSTAT3 activity and focused on monocytes, which are considered first-line immune sensors given their rapidity of response to immune perturbation, and so most likely to be sensitive to our time-limited study protocol(*33*). Moreover, catecholaminergic neurons of the sympathetic nervous system are present in the bone marrow and influence monocyte migration(*34*). The apparent significant reduction of peripheral monocyte pSTAT3 activity following modulation of IPL-PCC connectivity in RA patients not receiving JAKi therapy and otherwise on stable doses of immunotherapy is intriguing. While this may represent a specific and direct interference of JAK-STAT signalling, it could more simply mark out a general attenuation of the innate immune system. Indeed, VNS also appears to modulate pSTAT3 in the myeloid compartment, although the effect is indirect via engagement of α7 nicotinic acetylcholine receptors (α7nAChR)(*35*). Powered, multi-omic-based, clinical studies are required to follow up on this exploratory work to elucidate the precise immune mechanisms within both blood and tissue compartments. These will then directly inform more precise pre-clinical causal determination of the hierarchically dominant immune mechanisms. A similar reverse translation approach will enable definitive mapping of the interdependent brain-based circuitry.

Currently, we propose a neuro-immune framework building on our observed treatment-associated shifts in IPL–insula and DMN–insula coupling and the rTMS-induced change in IPL–PCC coupling and peripheral blood monocyte phosphorylation. In this framework, the IPL functions as an inflammation-responsive sensory region; the posterior insula indexes amplified signals of central sensitisation–like pain; the PCC integrates these signals within the DMN and coordinates adaptive homeostatic responses. In our previous work, higher peripheral inflammation was associated with more positive IPL connectivity to multiple brain networks(*8*), supporting the plausibility of an IPL “sensor” that tracks the immune state. More broadly, experimental immune activation can also alter brain function and recalibrate large-scale networks relevant to pain and self-processing(*36*). Within this pathway, the insula is positioned to receive peripheral sensory and visceral input and has been described as the primary interoceptive cortex(*37*). Consistent with this role, activity in the left posterior insula has been linked to pain intensity(*38*), and anatomical inputs of sensory neurons indicate that the insular cortex is a potential site of immune and interoceptive integration. Preclinical data revealed that projections from the peripheral source of inflammation and projections from the insula formed a nexus in two main autonomic outputs from the brainstem controlling parasympathetic and sympathetic outputs(*39*). Further, a landmark preclinical paper(*40*) demonstrated reactivation of specific neuronal ensembles in the insula, which were previously activated during a systemic peripheral inflammation stimulus, and induced inflammation at the original peripheral site. Taken together, these data implicate the insula at the crossroads of neuro-immune communication.

The PCC is highly connected anatomically and is a core DMN region, supporting its role as a hub that potentially links internally oriented representations with interoceptive and sensory signals(*41*). This integration could engage a homeostatic reflex arc through coupling with autonomic and arousal effector systems. In a recent 7T fMRI map of the human allostatic-interoceptive system, the PCC showed prominent connectivity with lower brainstem nuclei, including the locus coeruleus, parabrachial nucleus, and the medullary viscero-sensory-motor nuclei complex(*16*). In our exploratory analyses, left IPL rTMS altered PCC–locus coeruleus connectivity and left IPL–hypothalamus connectivity (Error! Reference source not found.) alongside changes in monocyte activity, which supports a role for DMN hubs in neuro-immune coupling. Both the locus coeruleus and hypothalamus have been implicated in DMN connectivity related to interoceptive signalling, and the hypothalamus region was defined in prior work linking its activity to TNF and ACTH after intravenous endotoxin(*17*). This circuit parallels the inflammatory reflex(*10*) in framing pain–inflammation coupling as a centrally integrated, reflex-like homeostatic loop in which neural output can inhibit cytokine release. A key difference is the control architecture: the inflammatory reflex is classically organised around vagal afferent–efferent signalling, whereas our model places integration in cortical DMN hubs with downstream engagement with autonomic effector nodes such as the hypothalamus and locus coeruleus, which drive sympathetic outflow and can regulate inflammation. These findings would need to be reverse-translated into mechanistic animal models to test whether an inflammation-sensitive IPL node and a pain-intensity–linked posterior insula node converge on PCC/DMN integration that then shapes autonomic output, pain experience, and peripheral inflammatory tone.

The underpowered, exploratory aspects of this study limit the interpretation of biological relationships with patient-reported outcomes. Future clinical studies should also adopt multiple regular stimulation sessions, as with the successful VNS trials. This is more likely to deliver the sustained immune effect that will be of greatest clinical relevance and help determine if the preliminary statistically significant improvement in pain NRS observed here can be replicated with a greater, more clinically meaningful size of effect and across different pain sub-types (e.g. nociplastic pain). Further limitations included the absence of a randomised design to enable robust comparison of the differential effect of JAKi with anti-TNF and placebo. Moreover, the follow-up period between the studies varied (JAKi,12w; anti-TNF/placebo, 8w), although the sub-group analysis of the filgotinib patients who undertook an interim scan at 4w indicates that the bottom-up effect mostly occurs within this short time window. Finally, these data are specific to RA and so are not generalisable to other inflammatory conditions. The underlying principle of a peripherally driven neuroplasticity subserving central sensitisation, however, also converges with data from osteoarthritis patients who demonstrate normalisation of distinct insula-based functional connectivity following joint replacement(*42*), potentially marking alternative, non-inflammatory, neural interfaces.

In summary, peripherally directed JAKi reverses neurobiological markers of central sensitisation in RA, specifically normalising IPL-insula functional connectivity. This provides the first human experimental medicine evidence for a role of systemic inflammation pathways in the sensitisation of central nociplastic pain pathways in patients with inflammatory disease and thus offers a biological rationale for the augmentation of anti-inflammatory therapies for the management of burdensome nociplastic pain in select RA patients. Such patients could be stratified according to dysfunction IPL-insula connectivity, a hypothesis which requires to be tested in a future clinical trial. Moreover, complementary exploratory data have informed a framework for a novel neuro-immune circuit intrinsic to central sensitisation that now demands deeper mechanistic investigation.

## MATERIALS AND METHODS

### Study design (Fig. 7)

#### JAKiDiscovery Cohort

This was a single-centre, observational test–retest study of adults with moderate to severe active RA scheduled to start filgotinib (JAK1 inhibitor) as part of standard care. Exclusion criteria included contraindications to MRI, major neurological disease, and prior exposure to targeted synthetic DMARDs for RA. Participants attended baseline and follow-up visits at weeks 4 (optional) and 12 after treatment initiation; each visit included 7T MRI, blood sampling, and clinical and behavioural assessments. We measured resting-state fMRI as well as EULAR Disease Activity Score (DAS28) and the Fibromyalgia degree – 2011 ACR survey criteria (a subjective score of nociplastic pain, range 0-31).

**Fig. 7.**
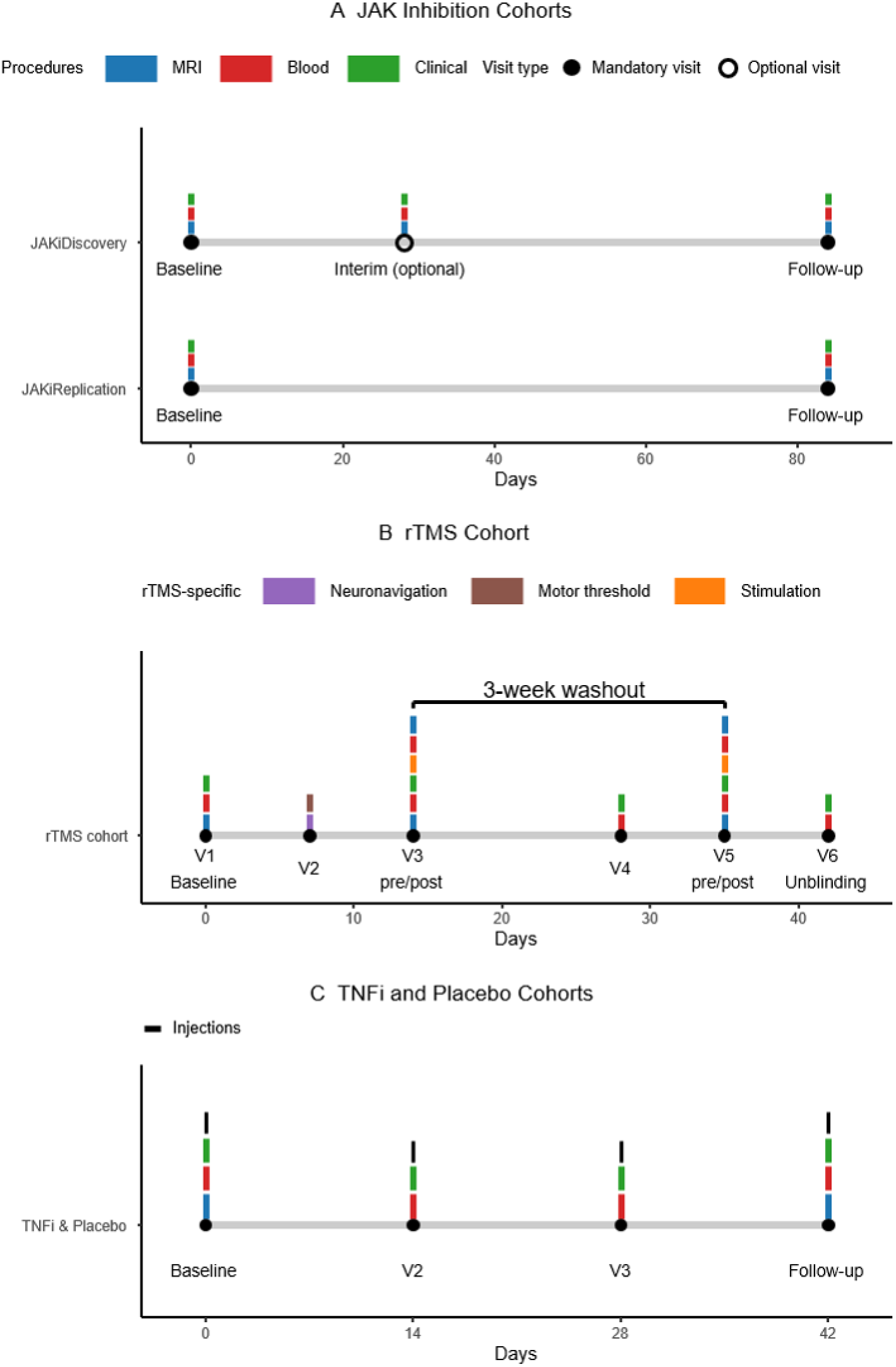
Integrated study schedules across cohorts. (A) JAK inhibition cohorts include JAKiDiscovery (with optional interim visit at week 4) and JAKiReplication. At each visit, procedures included MRI, blood sampling, and clinical/behavioural assessments (colour blocks). (B) rTMS crossover schedule with six visits, including neuronavigation and motor threshold assessment at V2; two stimulation sessions (V3 and V5) in pre- and post-stimulation measurements were taken. (C) Shared schedule for both arms (TNFi and placebo) with injections administered at days 0, 14, 28, and 42.

#### JAKiReplication Cohort

This was a single-centre, observational test–retest study of adults with RA scheduled to start baricitinib (JAK1/2 inhibitor) as part of standard care. Recruitment, key exclusions and data collection were as for the JAKiDiscovery cohort. Participants attended baseline and a week 12 follow-up visit.

#### TNFi and placebo Cohort

This was a two-centre (Glasgow and Cambridge) randomised study in adults with moderate to severe active RA scheduled to start adalimumab as part of standard care. Eligibility included self-reported sickness behaviour (fatigue, depression, or anxiety), with at least one component >4 on a Numerical Rating Scale (NRS). Key exclusions were prior biologic DMARD therapy, contraindications to MRI and major neurological disease. Participants underwent 7T MRI and assessments at baseline and at six weeks after four injections of adalimumab or placebo.

#### rTMS Cohort

This was a randomised, sham-controlled, crossover study in adults with active inflammatory RA and chronic widespread pain. Eligibility required CRP >6 mg/L or ESR >20 mm/hour and at least one swollen joint, with stable usual medication over the six-week study period. Exclusion criteria included contraindications to TMS (including seizure history), contraindications to MRI and major neurological disease. Participants received one active and one sham rTMS session targeting the left IPL, separated by three weeks, with 7T resting-state fMRI acquired before and after stimulation. Active rTMS consisted of a single 20-minute session delivered with a figure-of-eight coil at 10 Hz and 90% of resting motor threshold, for a total of 1200 pulses. Pulses were delivered as 20 trains of 6 s duration with 54 s intertrain intervals. This high-frequency protocol was chosen because previous studies show that parietal stimulation can modulate large-scale network activity and cortico-cortical connectivity(*43*), and may also produce analgesic effects in chronic pain, including fibromyalgia(*44*). Blood, as well as pain and fatigue NRS were taken before and after stimulation.

All participants in these studies gave full informed consent. Further details on eligibility criteria, visit schedules, randomisation, blinding procedures, clinical, and behavioural outcomes are provided in the Supplementary Materials.

### Peripheral immune blood assays

The pSTAT3 in monocytes plays a crucial role in regulating inflammatory immune responses. In RA, IL-6-mediated activation of the JAK1-STAT3 axis represents a central pathogenic pathway, with phosphorylated STAT3 promoting transcriptional cascades that sustain inflammation(*45-47*). Therapeutic interventions targeting this pathway, including IL-6 blockade and JAK inhibition, reduce STAT3 phosphorylation and are associated with reduced inflammation and clinical improvement(*48-50*). Accordingly, reduced inducible pSTAT3 reflects diminished responsiveness to IL-6-driven inflammatory signalling, which is consistent with an anti-inflammatory or immune-normalising effect.

We employed an optimised phospho-flowcytometry assay to assess changes in pSTAT3 levels in monocytes before and 1 hour after rTMS. Peripheral blood mononuclear cells (PBMCs) collected at the time of the study visits were frozen and stored in liquid nitrogen before processing. A total of 16 PBMCs samples from 8 participants with matched active and sham conditions were processed. PBMCs were thawed and stimulated with IL-6 at a final concentration of 100ng/µl, mimicking pro-inflammatory milieu. Cells were then stained for surface antigens using fluorescently conjugated antibodies, fixed and permeabilised, and subsequently stained for intracellular antibodies targeting pSTAT3; surface markers included: CD3, CD4, CD8, CD14, CD25, and CD127. This protocol was established and validated internally in our laboratory and has been applied successfully across several inflammatory diseases, ensuring methodological consistency and reliability. The protocol enabled profiling of dynamic IL-6-induced JAK1-STAT3 signalling responses in circulating monocytes following rTMS intervention.

### Power calculation

We acquired resting-state fMRI at 7T to increase statistical power for functional connectivity analyses. Relative to 3T, 7T can provide an increase in SNR, improving the spatial resolution and accuracy of functional network estimates(*51*). These gains translate to improved power to detect small, group-level effects, with published comparisons suggesting that about 25 participants at 7T can achieve power similar to about 38 participants at 3T(*52*).

The higher statistical power and within-subject design in JAKiDiscovery meant we expected to observe large effects. Using a Paired Samples T-test power calculation, a large effect size (Cohen’s d = 0.9) expected for our IPL-Insula marker, detectable with 80 % power at α=0.025, would require 15 participants. All other analyses were considered exploratory and were not part of the calculation.

### Neuroimaging analysis

Brain imaging data were collected on a 7T MAGNETOM Terra MRI scanner (Siemens Healthineers, Erlangen, Germany). The data included structural and resting-state fMRI scans. Details on resting-state fMRI data preprocessing are provided in Supplementary Materials and Methods. Patient data were excluded if more than 20% of their functional volumes were omitted (60 volumes) due to motion or were outlined as extreme outliers (mean – 3SD) due to low effective degrees of freedom for baseline or follow-up, as per instruction(*53*).

Both structural and functional scans were normalised into MNI space(*54*). Brain subregions of the insula were based on previous findings in patients with FM(*55*), whose connectivity was associated with pain thresholds, originally defined by Taylor et al.(*56*). The regions included were the anterior, middle, and posterior insula bilaterally. The seed regions were created as spheres (6-mm diameter) using the MarsBaR toolbox(*57*). The left IPL was based on coordinates (x = -50, y = -50, z = 40) from our previous paper, depicting associations with CRP and ESR and the ACR FM scale(*9*). The DMN definition was based on Yeo’s 7 parcellations of the brain into its main neural networks(*14*). Our previous work identified a left IPL-connected cluster spanning the left mid and posterior insula(*9*). Because the insula comprises anterior, mid, and posterior subdivisions with distinct anatomical and functional properties that differently associate with fibromyalgia(*55*), we parcellated this cluster into two prespecified targets, the left mid insula and left posterior insula. We also examined left IPL connectivity with the remaining four insula subregions as exploratory analyses (Error! Reference source not found.).

ROI-to-ROI analysis was then conducted on the pre-processed (without smoothing) and denoised data from these seeds, where connectivity was estimated between the left IPL and the left mid and left posterior insula subregions (FDR-corrected for the two comparisons). The analyses of the IPL and DMN with the other insula subregions were exploratory, so no multiple comparison correction was done.

The pre-processed data were also used for functional connectivity Multivariate Pattern Analysis (fc-MVPA) to identify regions whose pattern of connectivity with the rest of the brain significantly changed after treatment(*13*). Five eigenvectors were set up as per the instructions (5:1 ratio with the number of participants). All five eigenvectors were used in a within-subjects contrast to compare follow-up to baseline scans (omnibus F test), with false discovery rate (FDR) correction for multiple comparisons at p < 0.001.

### Statistical analyses

Comparisons between baseline and follow-up timepoints of neuroimaging, clinical, and flow cytometry variables were tested using paired-sample t-tests. Non-normal distributions based on the Shapiro-Wilk test used the non-parametric Wilcoxon signed-rank test instead. Linear mixed models were used in the rTMS cohort dataset to explore interaction effects between time (pre, post) and condition (active, sham). All statistical analyses were conducted in JASP 0.95(*58*).

## Data Availability

All data produced in the present study are available upon reasonable request to the authors.

## Acknowledgements

We would primarily like to thank the four cohorts of study participants. We also thank Vinod Kumar, Neil McKay, James Dale, Lindsay Robertson, Duncan Porter, and Stefan Siebert for their help with the recruitment of participants from their clinics. We also thank all the members of the radiography and clinical teams at the Queen Elizabeth University Hospital in Glasgow (Yvonne McLennan, Rosie Woodward, Laura Dymock, Fiona Savage, and Nicola Tynan) and the University of Cambridge (Neal Chauhan, Donna Abercrombie, Jane Rowlands, Thelma Mushapaidzi and Anne Meadows) for their role in data collection. We thank Jennifer Barrie, Richard Hohne, and Anna Grenz for processing the blood samples from the four studies. We also like to recognise Hanan Awan for his administrative support.

## Funding

The studies were sponsored by NHS Greater Glasgow and Clyde and funded by the following:

JAKiDiscovery: Alfa Sigma S.p.A. (TEMPO)

JAKiReplication: Lilly (SOAR GB-2011)

rTMS cohort: Arthritis UK (ProBEPP 13044)

TNF inhibitor and placebo cohort: Medical Research Council (REALISE MR/S012230/1). This study was also supported by CARE (Cambridge Arthritis Research Endeavour) and NIHR Cambridge Biomedical Research Centre (NIHR203312).

The views expressed are those of the authors and not necessarily those of the NIHR or the Department of Health and Social Care.

## Author contributions

All authors were involved in drafting the article or revising it critically for important intellectual content, and all authors approved the final version to be published.

Study conception and design: NB, KS, JTP, JC, DJC, CMK, AS, IBM, GT

Acquisition of data: KS, JTP, AM, JB, NA, AV, FS, MA, TL, NS, DP, GK, SW, GT

Analysis or interpretation of data: KS, NB, JC, JTP, FS, SW, AM, SA, CMK, AS, EI, LP, CG, NS, IBM, DJC, GT.

## Competing interests

Eli Lilly and Alfasigma have provided funding to N.B. for data acquisition. N.B. has consulted and served on speakers bureaus for Eli Lilly, Alfasigma and AbbVie. I.B.M has received research grant funding from Eli Lilly and UCB and consulted for AbbVie, Eli Lilly, Pfizer and UCB. N.S. has received research grant funding from Gilead, GSK, UCB, Kirin, and AbbVie and accepted fees for independent educational sessions from Alfasigma, AbbVie, Eli Lilly and Roche. M.A has consulted for Alphasigma. The remaining authors declare no competing interests.

## Data and materials availability

Anonymised raw imaging data will be made available to qualified researchers upon reasonable request through a material transfer agreement. All statistical analyses are available at: https://github.com/krisbg95/JAKiDMN.

## Notes

### Author Declarations

Ethical approval for this work was given by the South West - Cornwall & Plymouth Research Ethics Committee, the South West - Central Bristol Research Ethics Committee, the London Bridge Research Ethics Committee, and the London - Brent Research Ethics Committee.

