## Supplementary_Information for "JAK-STAT pathway inhibition modulates centrally sensitised default mode network hubs in rheumatoid arthritis pain"

### SUPPLEMENTARY MATERIALS

#### Recruitment, screening, and eligibility

JAKi cohorts (Discovery; Replication)

Participants were recruited from rheumatology clinics within NHS Scotland. Screening was performed by the research team using clinical interviews and review of the medical records.

Inclusion criteria: Participants, between 18 and 75 years of age, with moderate to severe active disease in line with local guidance for baricitinib (previous failure of at least 2 DMARDs)(59) and filgotinib(60).

Exclusion criteria: Contraindication to MRI; pregnancy or breastfeeding, severe physical impairment (for example, blindness, deafness, or paraplegia); major confounding neurological disease (for example, multiple sclerosis, stroke, or traumatic brain injury); prior exposure to targeted synthetic DMARDs for RA (for example, baricitinib or tofacitinib).

TNFi and placebo cohort (adalimumab versus placebo)

Participants were recruited from rheumatology outpatient clinics in NHS Greater Glasgow and Clyde and Cambridge University Hospitals NHS Foundation Trust.

Inclusion criteria: adults with active RA scheduled to start adalimumab as part of standard care; reported sickness behaviour (fatigue, depression, or anxiety), with at least 1 component greater than 4 on a numeric rating scale.

Exclusion criteria: prior biologic DMARD therapy; intramuscular or intra-articular steroid injection within 4 weeks before baseline; serious infection, including sepsis, tuberculosis, or opportunistic infections; contraindication to MRI; pregnancy or breastfeeding; severe physical impairment; major confounding neurological disease.

rTMS cohort

Participants were recruited from secondary care clinics in NHS Greater Glasgow and Clyde and NHS Lanarkshire. Eligibility was confirmed by a medically qualified member of the research team through medical record review.

Inclusion criteria: adults with active inflammatory RA and chronic widespread pain by ACR definition; willing to maintain usual medication over the 6-week study period; had objective evidence of active inflammation, defined as CRP greater than 6 mg/L or ESR greater than 20 mm/hour, and at least 1 swollen joint.

Exclusion criteria: contraindication to TMS, including a history of seizures; contraindication to MRI; pregnancy or breastfeeding; severe physical impairment; major confounding neurological disease.

#### Visit schedules and procedures

JAKiDiscovery: Visit 1 was baseline and included consent and demographic data collection. Visits 2, 3, and 4 were scheduled for days 14, 28, and 84 after treatment initiation, with an allowable window of plus or minus 7 days. Visits 2 and 3 were optional. Each visit included MRI, blood sampling, and clinical and behavioural assessments.

JAKiReplication: Visit 1 was baseline and included consent and demographic data collection. Visit 2 was scheduled for day 84 after treatment initiation, with an allowable window of plus or minus 7 days. Each visit included MRI, blood sampling, and the same clinical and behavioural assessments used in JAKiDiscovery.

rTMS cohort: This was a randomised, sham-controlled, crossover study with 2 stimulation sessions separated by 3 weeks. The study comprised 6 visits. Visit 1 included consent, medical history, confirmation of eligibility, disease activity assessment, and randomisation of stimulation order. Visit 1 also included a short MRI session to acquire a structural scan and to define the left IPL target for neuronavigation (Brainsight®). Visit 2 occurred 7 days after Visit 1 and included neuronavigation and motor threshold assessment. The assessment involved a single pulse TMS to stimulate the left motor cortex, which was located with the aid of neuronavigation. The motor threshold is the minimum intensity required to elicit a motor evoked potential in the contralateral hand in at least 50% of trials. Visit 3 occurred 14 days after Visit 1 and included an MRI scan before and after the first stimulation session, with pain and fatigue ratings recorded before and after stimulation. Visit 4 occurred 28 days after Visit 1 and included clinical and behavioural assessments only. Visit 5 occurred 21 days after Visit 3 and repeated the Visit 3 procedures with the alternate stimulation condition. Visit 6 occurred 7 days after Visit 5 and included unblinding and clinical and behavioural assessments.

TNF inhibitor and placebo cohort: Participants attended a baseline visit and a follow-up visit at 6 weeks after 4 injections of adalimumab or placebo. Each visit included MRI and clinical and behavioural assessments.

**Randomisation, allocation concealment, and blinding**

*JAKi cohorts (Discovery; Replication):* all participants received treatment with no control arm as part of the study, and consequently, no randomisation took place.

rTMS cohort: Participants were randomised to receive active rTMS first or sham first. Sham stimulation used a sham coil designed to mimic auditory and scalp sensations without intended brain modulation. The crossover interval between sessions was 3 weeks. Unblinding was performed after completion of the second stimulation period at Visit 6.

TNF inhibitor and placebo cohort: Participants were randomised in a 1:1 ratio to adalimumab or placebo. Randomisation was implemented through a central interactive voice response system. Randomisation was stratified by site and used randomly permuted blocks. The randomisation schedule was prepared without the involvement of investigators or the study statistician.

**Clinical and behavioural assessments**

The clinical evaluation included our outcomes of interest: (1) the 2011 ACR FM criteria(*61*), a proposed proxy measure of nociplastic pain previously validated in a RA cohort with fMRI(*7*). Total FM scores (0–31) were used as a continuous variable to express the degree of nociplastic pain (FMness)(*62*) while the PROMIS Depression 8a (short form, 8 questions) was used to estimate a depression score(*63*). Demographic data (age, sex, and BMI measured at the time of the visit), RA disease activity data in the form of EULAR Disease Activity Score (DAS28), which includes swollen and tender joint counts, CRP, and global VAS assessment, were collected at the time of the visit.

**MRI data collection**

Participants undertook scans on a 7 Tesla Siemens MAGNETOM Terra (Siemens, Erlangen, Germany) in Glasgow (UK) using a radiofrequency (RF) head coil with a single transmit channel and 32 receive channels. Scan protocols included whole-brain T1-weighted structural imaging using MP2RAGE (REF) (TR = 5000 ms, TE = 1.94 ms, inversion time (TI1) = 700 ms, TI2 = 2700ms, flip angle1 = 4°, flip angle2 = 5°, FOV = 240 mm, with 208 slices, 0.8 mm isotropic voxels) and whole-brain fMRI using a T2*-weighted, multiband EPI sequence (TR = 1500 ms, TE = 25 ms, flip angle = 65°, FOV = 192 mm, multiband slice acceleration factor = 4, in-plane acceleration GRAPPA = 2, 80 axial slices, interleaved, 307 volumes at 1.5 mm iso-voxel). An 8-minute resting-state fMRI scan was performed for the analysis, during which individuals were asked to lie supine in the scanner and keep their eyes open, focusing on a fixation cross without engaging in any specific task.

#### MRI data pre-processing

Preprocessing and later analysis of the images were carried out using SPM12 within the functional connectivity CONN toolbox 22.v2407(*64*), running in MATLAB R2023b. Preprocessing included the default MNI pipeline by CONN: realignment, slice-timing correction, ART-based motion outlier detection, coregistration, functional and structural segmentation, Montreal Neurological Institute (MNI152) template normalisation and 8-mm smoothing (convolution with an 8 mm full-width at half maximum Gaussian Kernel, not used for ROI-to-ROI analysis). All the scans were visually inspected for artefacts. Individual volumes were omitted from the analysis if they had over 0.9 millimetres of motion and a global BOLD signal of over five standard deviations. A patient was considered for exclusion from analysis if more than 20% of their functional volume was omitted (60 volumes). Signals from white matter and cerebrospinal fluid were extracted via the CompCor procedure (5 PCA components for each), and motion parameters (6 directions and first-order derivatives) were entered into the analysis as covariates of no interest via ordinary least squares regression. A bandpass filter (frequency window: 0.008–0.09 Hz) was applied to remove linear drifts and high-frequency noise from the data.

#### Serious adverse events

No serious adverse events were reported across all studied cohorts.

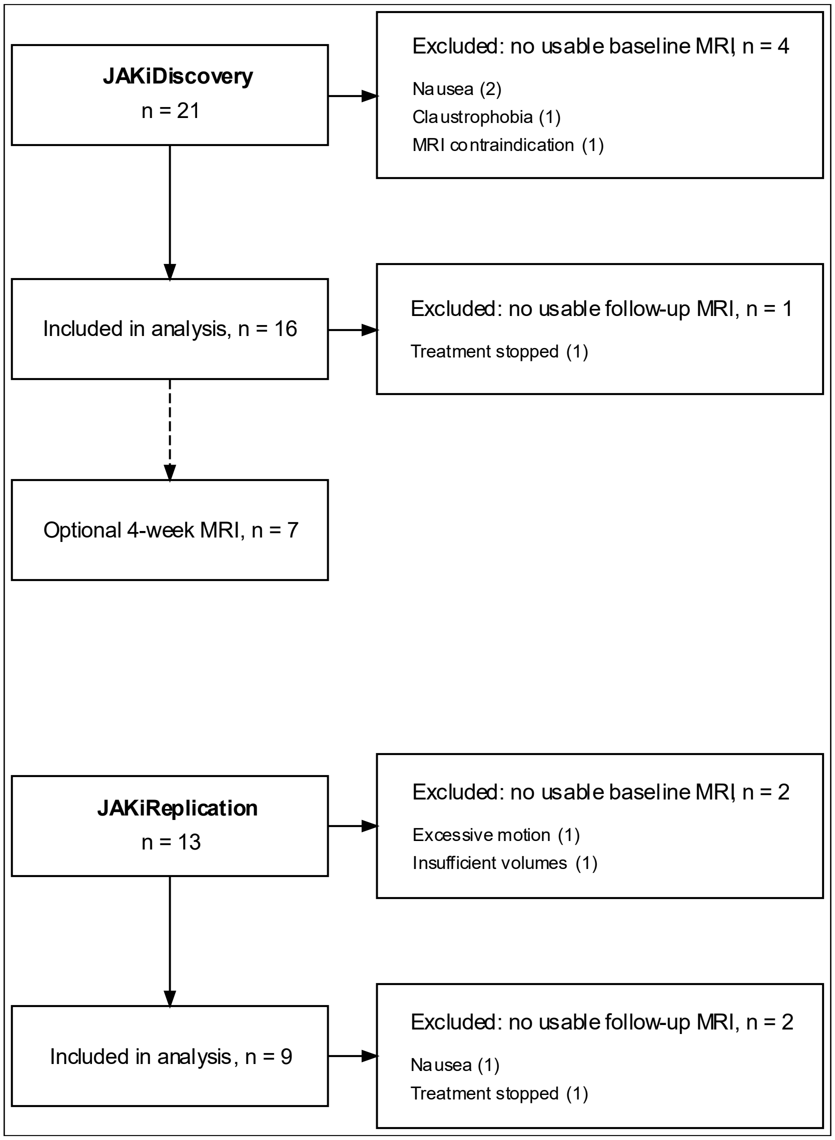


**Fig. S1. Recruitment and follow-up of the JAKiDiscovery and JAKiReplication cohorts.**
Participants were included in the MRI analysis only when both baseline and 12-week follow-up scans were available and of adequate quality. Exclusions are categorised according to whether they occurred because of unusable or missing baseline MRI data or unusable or missing follow-up MRI data, with individual reasons shown for each cohort.

| | | | t | df | p |
|---|---|---|---|---|---|
| LantIC-Left IPL Baseline | - | LantIC-Left IPL Follow-up | -0.15 | 15 | .88 |
| LmidIC-Left IPL Baseline | - | LmidIC-Left IPL Follow-up | 0.76 | 15 | .46 |
| **LpostIC-Left IPL Baseline** | **-** | **LpostIC-Left IPL Follow-up** | **-2.56** | **15** | **.02** |
| RantIC-Left IPL Baseline | - | RantIC-Left IPL Follow-up | -0.31 | 15 | .76 |
| RmidIC-Left IPL Baseline | - | RmidIC-Left IPL Follow-up | -0.66 | 15 | .52 |
| RpostIC-Left IPL Baseline | - | RpostIC-Left IPL Follow-up | -1.19 | 15 | .25 |

**Table S1. Changes in IPL-insula connectivity in JAKiReplication (Filgotinib).**

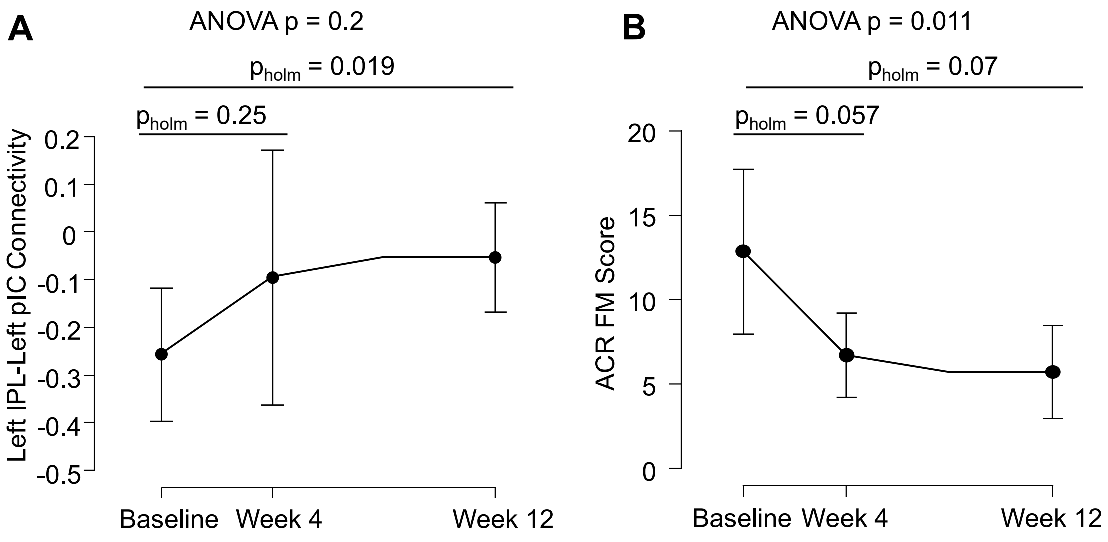


**Fig. S2. Filgotinib demonstrates swift normalisation of connectivity and symptoms using interim data.**

(A) Seven patients completed MRI and ACR FM assessments at baseline, 4 weeks, and 12 weeks during filgotinib treatment. Repeated-measures ANOVA did not show a significant time effect on left IPL–left posterior insular cortex (pIC) connectivity ($F$ (2,12) = 1.85, p = 0.2), but mean connectivity shifted toward zero from −0.257 (SD = 0.10) at baseline to −0.095 (SD = 0.26) at week 4 and −0.054 (SD = 0.20) at week 12. (B) ACR FM scores followed a similar pattern, decreasing from 12.86 (SD = 5.98) at baseline to 6.71 (SD = 4.39) at week 4 and 5.71 (SD = 3.40) at week 12. The line plots display the mean values and 95% confidence intervals (normalised). Post hoc t-tests are displayed with their Holm-corrected p-values.

| | | Frequency Female | Percent Female | Frequency Male | Percent Male |
|---|---|---|---|---|---|
| Gender | anti-TNF | 15 | 71.4% | 6 | 28.6% |
| Gender | Placebo | 3 | 33.3% | 6 | 66.7% |
| | | Median | Mean | Std. Deviation | IQR |
| Age | anti-TNF | 61.000 | 59.286 | 10.932 | 14.000 |
| Age | Placebo | 49.000 | 52.111 | 12.850 | 16.000 |
| Disease Duration | anti-TNF | 6.333 | 7.712 | 6.427 | 7.667 |
| Disease Duration | Placebo | 3.500 | 7.176 | 9.552 | 6.917 |
| DAS28 Baseline | anti-TNF | 4.910 | 4.620 | 1.219 | 1.520 |
| DAS28 Baseline | Placebo | 4.185 | 4.155 | 1.526 | 2.032 |
| SJC Baseline | anti-TNF | 4.000 | 6.048 | 6.515 | 5.000 |
| SJC Baseline | Placebo | 4.000 | 4.889 | 4.961 | 3.000 |
| TJC Baseline | anti-TNF | 11.000 | 10.000 | 8.620 | 13.000 |
| TJC Baseline | Placebo | 5.000 | 8.000 | 7.263 | 4.000 |
| CRP Baseline | anti-TNF | 7.000 | 14.143 | 16.206 | 11.000 |
| CRP Baseline | Placebo | 5.000 | 18.111 | 37.120 | 9.000 |
| ACR FM score Baseline | anti-TNF | 20.000 | 18.800 | 9.540 | 13.750 |
| ACR FM score Baseline | Placebo | 19.500 | 17.750 | 8.681 | 10.250 |
| Pain NRS Baseline | anti-TNF | 6.000 | 5.571 | 2.942 | 4.000 |
| Pain NRS Baseline | Placebo | 7.000 | 6.222 | 1.716 | 1.000 |

**Table S2. Baseline characteristics of the TNF inhibitor and placebo cohort.**

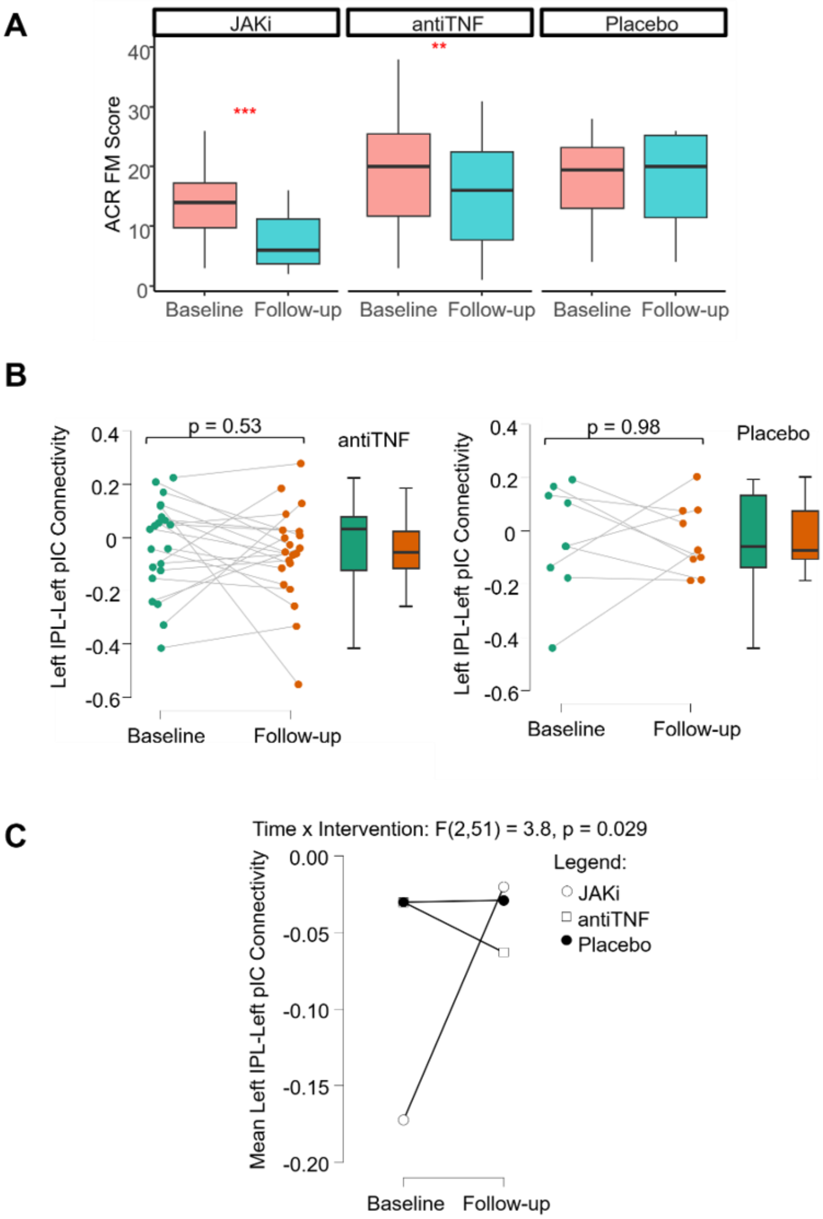


**Fig. S3. JAKi, anti-TNF and placebo effects on patient-reported nociplastic pain and bottom-up connectivity.** (A) Immunological therapies reduced patient-reported nociplastic pain but not placebo. The boxplots depict that JAK inhibition (filgotinib and baricitinib) and anti-TNF significantly reduced ACR fibromyalgia scores, but not placebo. *** $p < 0.001$, ** $p < 0.01$. (B) Bottom-up nociplastic pain functional connectivity changed in JAK inhibition but not in TNF inhibition or placebo. The boxplots depict the lack of change (paired t-tests) after anti-TNF and placebo in connectivity between the left inferior parietal lobule (IPL) and the left posterior insular cortex (pIC). (C) A significant interaction between time and type of intervention (repeated measures ANOVA) on mean left IPL-left pIC connectivity.

| | | | t | df | p |
|---|---|---|---|---|---|
| LantIC-DMN Baseline | - | LantIC-DMN Follow-up | -0.64 | 23 | .53 |
| LmidIC-DMN Baseline | - | LmidIC-DMN Follow-up | -0.47 | 23 | .64 |
| LpostIC-DMN Baseline | - | LpostIC-DMN Follow-up | -1.07 | 23 | .29 |
| **RantIC-DMN Baseline** | **-** | **RantIC-DMN Follow-up** | **-2.40** | **23** | **.02** |
| RmidIC-DMN Baseline | - | RmidIC-DMN Follow-up | -0.78 | 23 | .44 |
| RpostIC-DMN Baseline | - | RpostIC-DMN Follow-up | -0.26 | 23 | .79 |

**Table S3. Changes in DMN-Insula connectivity after filgotinib and baricitinib. Based on paired-samples t-tests between baseline and follow-up functional connectivity data.**

| | | Frequency Female | Percent Female | Frequency Male | Percent Male |
|---|---|---|---|---|---|
| Gender | JAKiD | 12 | 75% | 4 | 25% |
| | JAKiR | 5 | 55.6% | 4 | 44.4% |
| | | Median | Mean | SD | IQR |
| Age | JAKiD | 59.50 | 56.63 | 9.64 | 10.50 |
| | JAKiR | 55.00 | 55.78 | 10.28 | 11.00 |
| Disease Duration | JAKiD | 8.25 | 12.75 | 10.69 | 14.33 |
| | JAKiR | 11.17 | 12.46 | 9.81 | 10.29 |
| DAS28 | JAKiD | 4.54 | 4.64 | 1.45 | 2.01 |
| | JAKiR | 4.84 | 4.60 | 1.42 | 1.71 |
| SJC | JAKiD | 5.00 | 8.13 | 7.43 | 11.25 |
| | JAKiR | 7.00 | 7.22 | 3.56 | 5.00 |
| TJC | JAKiD | 9.00 | 11.06 | 8.98 | 14.50 |
| | JAKiR | 8.00 | 9.78 | 8.57 | 10.00 |
| CRP | JAKiD | 4.00 | 7.25 | 8.64 | 10.00 |
| | JAKiR | 2.00 | 11.22 | 21.40 | 10.00 |
| ACR FM score | JAKiD | 15.50 | 14.38 | 5.63 | 7.75 |
| | JAKiR | 12.00 | 13.67 | 5.64 | 4.00 |
| Pain NRS | JAKiD | 5.95 | 5.14 | 1.97 | 3.33 |
| | JAKiR | 5.00 | 5.59 | 2.75 | 3.50 |

**Table S4. Clinical profile of rheumatoid arthritis patients in the JAKiDiscovery (JAKiD) and JAKiReplication (JAKiR) cohorts before they received filgotinib or baricitinib, respectively.**

| | | Mean Change (SD) | Statistic (t) | p |
|---|---|---|---|---|
| DAS28 | JAKiD | -1.51 (1.49) | -4.06 | 0.001 |
| | JAKiR | -1.18 (1.5) | -2.37 | 0.043 |
| SJC | JAKiD | -1.94 (9.7) | -0.8 | 0.437 |
| | JAKiR | -5.33 (4.58) | -3.49 | 0.008 |
| TJC | JAKiD | -7.06 (7.51) | -3.76 | 0.002 |
| | JAKiR | -3.22 (4.58) | -2.11 | 0.068 |
| CRP* | JAKiD | -2 (4.5) | -1.08 | 0.292 |
| | JAKiR | -1 (2) | -0.84 | 0.44 |
| ACR FM score | JAKiD | -6.44 (6.52) | -3.95 | 0.001 |
| | JAKiR | -3.78 (5.24) | -2.16 | 0.062 |
| Pain NRS | JAKiD | -2.81 (2.37) | -4.42 | < 0.001 |
| | JAKiR | -2.72 (2.78) | -2.49 | 0.019 |

**Table S5. Changes in clinical profile after treatment in the JAKiDiscovery (JAKiD) and JAKiReplication (JAKiR) cohorts.**

* CRP had a non-normal distribution, so the table shows its median change (IQR) and the result of a Wilcoxon signed-rank test instead of a Student's t-test with a reported z-statistic.

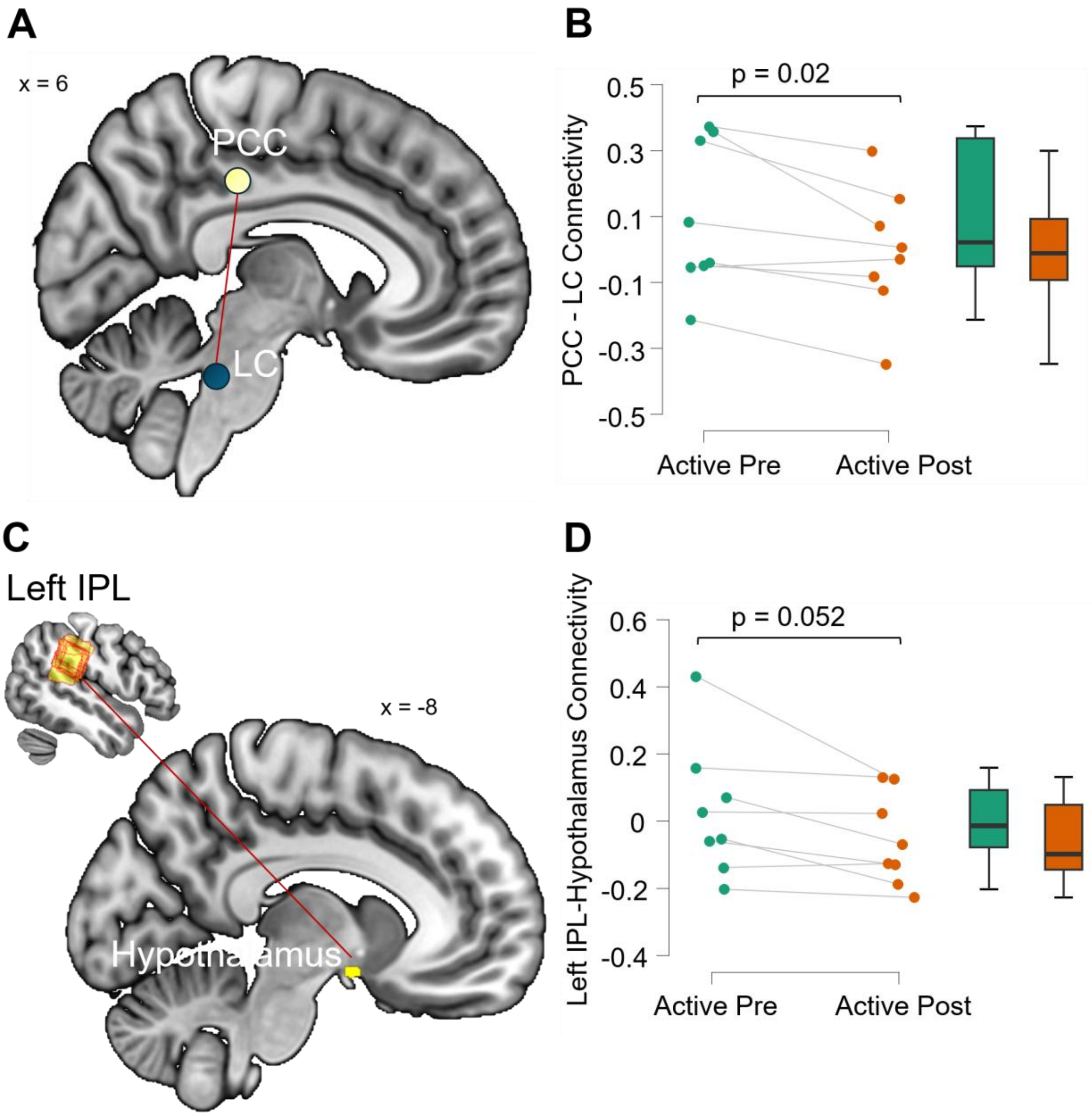


**Fig. S4. Active rTMS changes connectivity between the PCC and LC and between the left IPL and Hypothalamus.**

(**A**) The coordinates for the posterior cingulate cortex (PCC, MNI = -8, -34, 36) based on the fc-MVPA analysis in the JAKi cohorts, the Locus coeruleus (LC) based on a 7T allostatic–interoceptive system(*16*). (**B**) the single values and boxplots for the PCC-LC connection, paired t-test and respective p value demonstrating the effect of rTMS. (**C**) the left inferior parietal lobule (IPL) based on the individual coordinates used for rTMS, and the hypothalamus (MNI = -8, 0, -13), based on activity related to an inflammatory response(*17*). (**D**) the single values and boxplots for the left IPL-Hypothalamus connection and paired t-test results.
